# Changes in risk factor status and risk of future ASCVD events in primary prevention

**DOI:** 10.1101/2024.01.08.24300916

**Authors:** In-Chang Hwang, Chee Hae Kim, Jae-Young Kim, Jiesuck Park, Hye Jung Choi, Hong-Mi Choi, Yeonyee E. Yoon, Goo-Yeong Cho

## Abstract

Primary cardiovascular prevention highlights the management of risk factors through lifestyle changes and therapeutic interventions. While most previous studies focused on the presence of risk factors assessed only at baseline, temporal changes in the risk factor status were not considered. We aimed to evaluate the prognostic impact of temporal changes in risk factor status in primary prevention settings. A population-based cohort of 211,077 individuals who underwent repetitive 10-year atherosclerotic cardiovascular disease (ASCVD) risk assessments over 4–5 years, was identified from National Health Insurance Services claims data of South Korea. Changes in risk-factor status for blood pressure, glycemic control, cholesterol levels, smoking status, body-mass index, and physical activity were assessed. Hazard ratios (HR) for ASCVD events after follow-up screening were calculated based on increased or decreased risk factors between baseline and follow-up. Regardless of baseline status, increased risk factors correlated with higher future ASCVD risk, while a decrease corresponded to a lower risk. Notably, individuals initially without risk factors but developing three at follow-up faced significantly higher ASCVD risk (adjusted HR 2.74, 95% CI: 1.31– 5.76). Conversely, those decreasing three risk factors from 5 or 6 exhibited almost a 50% reduction in ASCVD risk. These findings show that primary prevention effects can be reflected by changes in risk factor status, underscoring the need to monitor temporal changes in risk factors for effective primary prevention.

## Main text

In clinical practice, primary prevention targets cardiovascular risk factors through lifestyle changes and therapeutic interventions, which can alter risk factor status over time and affect prognosis.(1) While prior studies typically assessed risk factors only at baseline, neglecting temporal changes,(2, 3) our recent publication demonstrated a significant correlation between the rate of change in 10-year ASCVD risk (Δ10-year ASCVD risk/year) and future ASCVD events, irrespective of baseline risk profiles.(4) Although changes in the 10-year ASCVD risk reflect overall risk management, real-world practice more prioritizes on controlling individual risk factors such as hypertension, diabetes, dyslipidemia, smoking status, physical activity, and obesity. Monitoring the occurrence or resolution of these risk factors is more intuitive for both physicians and patients than tracking changes in the 10-year ASCVD risk scores. Therefore, we aimed to assess the prognostic impact of temporal changes in risk factor status in primary prevention settings.

Data from the National Health Insurance Services (NHIS) Health Screening Cohort of South Korea, a nationwide claims database, were used.(4, 5) This study was approved by the institutional review board of Seoul National University Bundang Hospital and adhered to the 2013 revised Declaration of Helsinki. Among 1,108,369 individuals who participated in the screening program between 2009 and 2015, we excluded those without repeated screenings at a 4–5 year interval and those without sufficient data to assess changes in risk factors. Additionally, individuals with established ASCVD, malignancy, or end-stage renal disease were excluded. A total of 211,077 individuals were analyzed. The primary outcome was ASCVD events (myocardial infarction, acute coronary syndrome, stable or unstable angina, coronary revascularization, stroke, transient ischemic attack, or peripheral arterial disease) occurring after follow-up screening (4–5 years post baseline screening, until 2019). Changes in risk-factor status between baseline and follow-up screenings were assessed for 6 components: blood pressure (≥140/90 mmHg), glycemic control (fasting glucose of ≥130 mg/dL for diabetic patients; occurrence of diabetes for non-diabetic patients), low-density lipoprotein cholesterol level (≥160 mg/dL), smoking status (current smoking), body-mass index (≥25 mg/kg^2^), and physical activity. Appropriate physical activity was defined as at least 150 min of moderate-intensity exercise or 75 min of weekly vigorous-intensity exercise.(1) To determine the resolution of specific diseases (e.g., hypertension, diabetes, dyslipidemia), we examined claims for diagnostic codes and medications at follow-up screenings. Patients initially diagnosed but lacking these indicators at follow-up prompted further assessment of blood pressure, fasting glucose, and lipid profiles. Resolution was confirmed when diagnostic codes, medications, and screening measurements were all negative at follow-up. Study population was stratified based on the number of risk factors at baseline. Hazard ratios (HR) for ASCVD events occurring after the follow-up screenings were calculated according to changes in the number of risk factors between baseline and follow-up screenings. Multivariable Cox proportional hazard regression model was used, adjusting for age, sex, smoking status, presence of diabetes, total cholesterol, high-density lipoprotein cholesterol, systolic blood pressure, and treatment for hypertension. For subgroups with small number of patients, Cox proportional hazard models with Firth’s penalized likelihood procedures were applied. For patients initially presenting with 0, 1, or 2 risk factors, HR was calculated based on the increase in risk factors, using those with the lowest number of risk factors at follow-up as the reference group. Conversely, for patients with 3, 4, or 5–6 risk factors at baseline, HR was calculated based on a decrease in risk factors, with those having the highest number of risk factors at follow-up as the reference group.

Regardless of baseline risk factor status, an increase in risk factors correlated with higher ASCVD event risk, while a decrease corresponded to lower risk (**Figure**). Among individuals without any risk factors at baseline, those with 3 risk factors at follow-up had a higher risk of future ASCVD events (adjusted HR 2.74, 95% confidence interval: 1.31–5.76), compared to those with no change in risk factor status (no risk factors at both baseline and follow-up examinations). Similar trend was observed among individuals with poor risk factor control at baseline. Compared to those with 5 or 6 risk factors at both baseline and follow-up examinations, individuals with a decrease of 3 risk factors had reduced risk of ASCVD events by nearly 50%.

**Figure.**
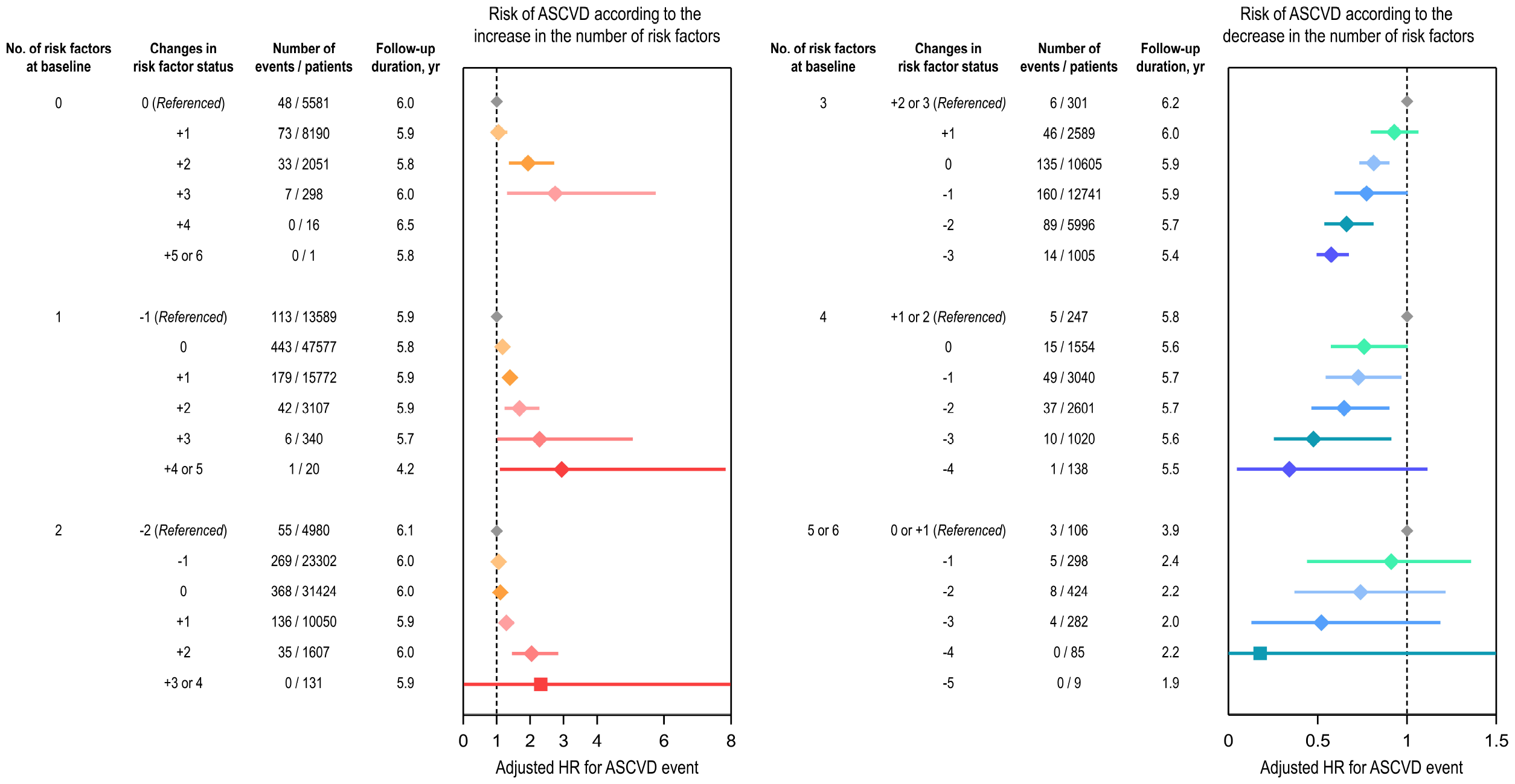
Associations between changes in risk-factor status and ASCVD events. The risk of ASCVD events is shown according to the increase (*left*) and decrease (*right*) in the number of risk factors between baseline and follow-up health screenings. Hazard ratios (HR) with 95% confidence intervals, calculated using multivariable Cox proportional hazard regression models, are depicted by rhombus marks (⍰) for conventional calculation and square marks (▪) for Firth’s penalized likelihood procedures. ASCVD, atherosclerotic cardiovascular disease; HR, hazard ratio; yr, year

Several limitations should be acknowledged in this study. Firstly, we relied on a claims database within a population-based cohort, potentially introducing selection bias despite the extensive coverage of the NHIS.(4, 5) Secondly, we did not assess the impact of medications in our analyses. Thirdly, small sample sizes with rare events in certain subgroups limited statistical significance, even with Firth’s penalized likelihood procedures. Finally, it’s important to note that each risk factor may carry a different prognostic weight, introducing limitations in our approach.

Nevertheless, this is the first population-based cohort study focusing on the temporal changes in cardiovascular risk factors in primary preventive setting. Unlike calculating changes in 10-year ASCVD risk estimates,(4) this approach provides practical evidence supporting risk factor control. Specifically, we demonstrated that primary prevention effects can be reflected by changes in risk factor status. Among individuals with appropriate risk factor control at baseline, newly developed risk factors significantly increase ASCVD events risk. Conversely, even in individuals with poor risk factor control at baseline, appropriate management can significantly improve prognosis. These findings emphasize the pivotal prognostic role of managing each risk factor in real-world practice.

## Data Availability

Anonymized data are publicly available from the National Health Insurance Sharing Service and can be accessed at https://nhiss.nhis.or.kr/bd/ab/bdaba000eng.do.

## Acknowledgments

We thank Professor Jeong-Ju Yoo, MD, PhD, for her dedication and support of this study.

